# Comparing the results of the application of moving and stationary sinusoidal gratings in the functionally assisted treatment of meridional amblyopia

**DOI:** 10.1101/2021.03.29.21251808

**Authors:** Uwe Kämpf, Svetlana Rychkova, Felix Muchamedjarow, Evelyn Heim

**Affiliations:** Caterna Vision GmbH; Svetlana Rychkova, Russian Academy of Sciences - Kharkevich Institute, Bolshoy Karetny per. 19, RF 127051 Moscow; Amblyocation GmbH

## Abstract

**Background:** The first approach to a frequency-selective visual training in amblyopia had been proposed in the sixties by the physiologist Fergus Campbell. During the stimulation, his grating was moving very slowly, i.e., only once a minute around its axis. Therefore, the CAM-stimulator was based exclusively on the proposed influence of spatial frequency selectivity, however there was no significant contribution of the temporal frequency of the stimulus. Accordingly, we are convinced that this was one of the main reasons for the failure of Campbell’s approach after the evaluation by a multiple of placebo-controlled studies. The aim of the present work, therefore, was to investigate in the influence of visual exercises, which contained moving gratings as compared to such of stationary gratings implemented into computer games on the development of visual acuity. Especially we were interested in the effects of such type of visual exercises, termed Focal Ambient Visual Acuity Stimulation (FAVAS), on the visual acuity development in patients with meridional amblyopia. We assume, in particular, that such a ring-shaped stimulus can reach all angular positions equally, and thus excite the most ametropic meridians, as compared to the least ametropic meridians, in a particular way.

**Methods:** Overall, we investigated in two groups of patients with meridional amblyopia who had been selected according to the age structure and to their type of disturbance. Using a cross-over design, the first group was alternately exercised 10 days with the moving followed by 10 days with the stationary grating stimulus, and the second group was alternately exercised 10 days with the stationary followed by 10 days with the moving grating stimulus, i.e. in reverse order. For the treatment, a sinusoidally modulated drifting grating had been implemented as a background stimulus into simple computer games. These games served for the attraction of attention of the children. For the present study these exercises were applied in the following two versions. The first version contained a concentrically outward-moving ring-shaped sinusoidal grating behind of the computer games during the process of visual stimulation. The grating was fixed at a spatial frequency of 0.3 cyc/deg with the time frequency of 1 cyc/sec, which gave an angular velocity of 3.33 deg/sec. The second version of the program contained the same grating, but in a non-moving stationary presentation. In order to additionally distinguish between the envisaged meridional stimulus effects, we examined our patients with regard to their corrected visual acuity using a directionally sensitive set of optical characters developed at the Kharkevich Institute. The monocular corrected visual acuity was tested in all patients at 4 meridians: 0 °, 45 °, 90 ° and 135 °. Additionally, in all patients of the first group and all patients of the second group, the binocular visual acuity was examined too.

**Results:** In the measurements of the corrected visual acuity along four meridians, a statistically significant improvement was found with alignment of the directional optical characters close to the meridian with maximal ametropia, and the minimal improvement in the orientation perpendicular thereto. In the patients of the both groups, the corrected visual acuity had significantly increased as a result of the treatment performed in the stage with a series of exercises with the moving sinusoidal grating. After the stage of treatment using the stationary grating, however, there was found no statistically significant improvement.

**Conclusions:** Exercises using FAVAS computer games that contained a moving sinusoidal grating resulted in a statistically significant positive dynamics of visual acuity development in the most ametropic meridian as compared to the least ametropic meridian. No statistically significant improvement was observed after exercises with the stationary grating.

## Introduction

Amblyopia is one of the most common ophthalmological disorder in young patients with a prevalence of 5-6 % [12]. While affected children are impacted in their daily activities and future job selection, it also increases the risk of a severe trauma for the better eye [32]. Since Sattler [44] re-introduced into the practice of applied strabology the method of occlusion, i.e. the patching of the fellow sound eye, was indisputably accepted as the gold standard of modern amblyopia therapy. Starting the occlusion early and adhering to it persistently, are two important factors that influence the effectiveness of this treatment. It has been shown that delayed diagnosis and treatment results in poor outcome [6].

On the other hand, notwithstanding that therapeutic strategies such as patching have been established in clinical practice for a long time, their success has been limited with a high rate of patients being resistant to therapy or not reaching normal visual acuity. Despite of an average occlusion therapy of more than two years, only one third of the patients achieved a complete visual acuity recovery and approximately one third still had residual amblyopia [3]. In view of this, pleoptics, as a system of visual exercises and stimulation methods in support of the standard occlusion procedure, had been developed early as a complementary treatment of amblyopia [4, 9, 11, 37, 38]. Although the concept of pleoptics is well known and established for many decades, this approach was not only complicated, time-consuming, but also requiring well-trained specialists; in sum, pleoptics requires an insupportably high expenditure of face-to-face therapy hours for patients and medical staff.

Recently, a paradigm shift in the therapeutic approach is aiming not at merely occluding the better eye, but rather additionally stimulating the amblyopic eye in line with pleoptic tradition. This is achieved with the aid of computer games, behind which there is a therapeutic stimulation concept and which thus make it possible that patients carry out pleoptic training online at home in combination with occlusion. Notably, a therapeutic software-based visual stimulation system for the complementary treatment of amblyopia had been developed two decades ago [21, 22, 23, 30, 35] by an interdisciplinary team of the Dresden University’s ophthalmologic clinics (Theo Seiler), the department of psychology (Uwe Kämpf), the faculty of informatics (Wilfried Mascolus) and including scientific partners (UKE Hamburg, Wolfgang Haase). It was shown that such visual stimulation training is of value as a complementary method in general [21] as well as for the treatment of therapy-resistant cases, for example, of residual amblyopia [23].

The software-based games for visual training are based on a special *focal-ambient visual acuity stimulation* (FAVAS). In the foreground of the screen, a *focal* computer game demands sensory-motor coordination, visual fixation performance and adherence from the children. Thus, the gaming activity serves for attention binding, which had been previously proven to be a decisive factor for the success of visual training exercises (Figure. 1). At the same time, an *ambient* stimulation is provided in the game’s background by a drifting sinusoidally contrast-modulated grating pattern of constant spatial and temporal frequency. Due to such periodicity, the drifting grating stimulus is assumed to induce resonance within and between filter systems of band-pass selective visual transmission channels.

**Figure 1:**
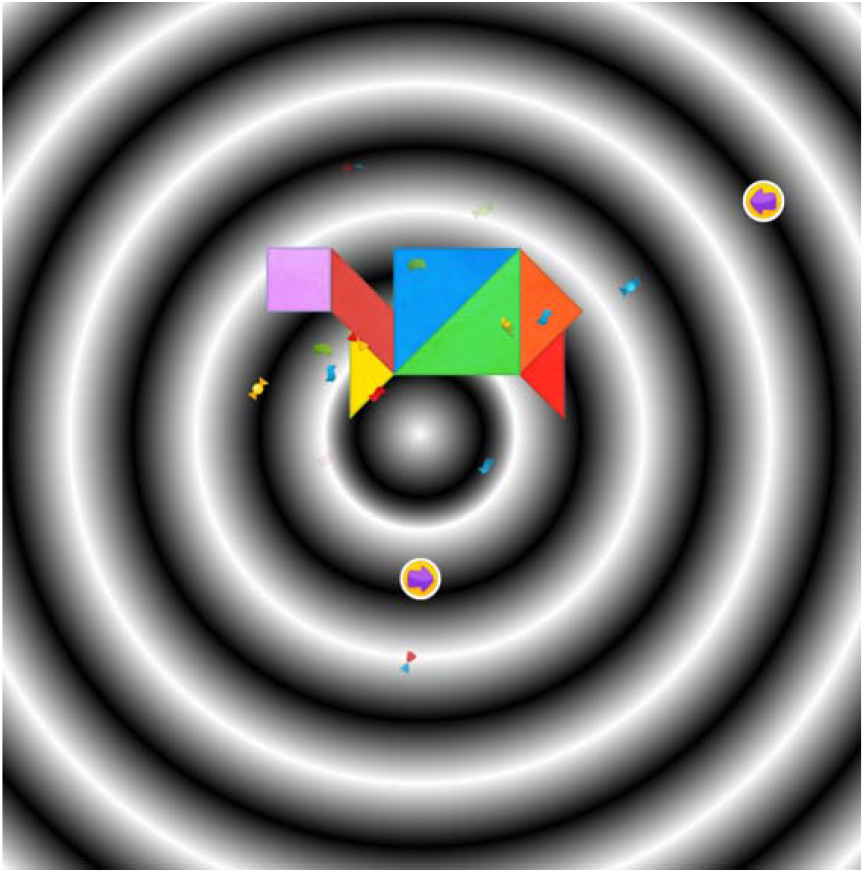
Example of FAVAS Stimulation in the backround and finished game for adherence in foreground.

This is relevant since in amblyopia the transmission quality of the *focal* system, which seems to predominantly filter the frequency-modulated correlates of form perception, appears to be more impaired than that of the complementary *ambient* system for the transmission of spectrally filtered motion correlates (cf. our discussion section of this paper for more details). Thus, at surface, the amblyopic vision disorder manifests itself in a focal dysfunction of the form channels of high spatial vs. low temporal frequency resolution [19]. Comparatively less disordered appears to be the functioning of ambient motion channels of low spatial vs. high temporal frequency resolution [40]. In light of this, our approach of focal-ambient visual stimulation (FAVAS) is designed to affect the disturbed focal form channels not directly but rather collaterally using the cooperative interplay with ambient motion channels [21, 22, 23].

## Objectives

This cooperative-synergy approach is crucial for FAVAS, since independently and differing from the above presented approach, the reference to frequency-bandwidth selective visual channels had been already the topic of a very early proposal of supportive amblyopia treatment by grating stimulation. The so-called CAM-stimulator had been developed by the British physiologist Campbell and his team in Cambridge [9]. Their results were later controversially discussed in view of placebo controlled trials [15, 27, 28, 31, 34]: Because of the unsuccessful replication studies, this attempt was empirically evaluated as a failure.

The Cambridge stimulator aimed at a direct stimulation of form channels, thus, omitting the possible synergy of the motion channels. His grating stimulus extremely slowly turned clockwise around the center of stimulation, i.e., in a nearly stationary or steady state way only once per minute around its axis. Therefore, the CAM stimulator was presumably solely based on the proposed influence of spatial frequency selectivity, so there was no significant contribution from the temporal frequency of the stimulus. Accordingly, we proved the question whether this critical feature might have been even the reason for the failure of Campbell’s approach which was shown as a result of the above-cited placebo-controlled studies.

In light of this, the main objective of our present investigation was to evaluate the influence of visual exercises containing moving gratings as compared to those of stationary gratings implemented in the background of computer games on the development of visual acuity. Thus, our primary outcome consisted in the investigation of the influence of visual exercises using moving gratings as compared to stationary gratings on the development of visual acuity. Pre-specifying from previous studies [21, 22, 23], we assume that the initial combination of FAVAS with occlusion as first-line therapy will increase the best-corrected visual acuity by at least two lines in a maximum of ten days in order to show the superiority over FAVAS as second-line therapy in a crossover design.

Another objective of the present study was to evaluate a possibly differential effect of this treatment in cases of so-called meridional amblyopia. As has been recently found, over 90% of amblyopic patients show an astigmatism > 0.5 dpt [48]. Consequently, meridional amblyopia often arises under astigmatism due to the visual-acuity difference between the differentially affected meridians. In patients with complex visual disorders, it may occur in combination with other forms of functional weakness, such as dysbinocular, anisometropic, or obscuration-related forms. It can also make a significant contribution to the reduction of visual performance in conditions of ocular fundus pathology.

In the present trial, as a secondary outcome, we were to ask for possible differences between the most affected (highest-ametropic) vs. the less affected (lowest-ametropic) meridian in their response to the FAVAS visual exercises in case of meridional amblyopia. Our hypothesis: FAVAS moving-grating stimulation combined with computer gaming and occlusion is more effective than stationary-grating sham stimulation combined with computer gaming and occlusion in treating children with meridional amblyopia,

According to our above declared objectives we aimed, in sum, at the following both outcomes. Primary outcome: best-corrected visual acuity (VA) after 10 days of stationary gratings (Sham-FAVAS) vs. 10 days of moving gratings (FAVAS). Secondary outcome: best-corrected visual acuity (VA) monocular in highest-ametropic vs. lowest-ametropic meridian. No changes to trial outcomes after the trial commenced were introduced in the a priori design.

## Patients and methods

### Patients

This prospective study adhered to the declaration of Helsinki, was conducted at a single center and was approved by the local ethics committee (05/2015, trial registration DRKS00022791). For this study an informed consent was obtained from patients’ legal guardians to take part in the present study, as well as to have the results of this research work published.

Eligibility criteria for participants were prescribed as follows: Sex of the children (male or female), their age (between 9 to 14 years old) and the type and impact of amblyopia. As a criterion for the impact we referred to the best corrected visual acuity in each amblyopic eye <0.8 measured by single optical characters. With respect to the type of, patients suffering from amblyopia ex anopsia with astigmatism were chosen, regarding our partial goal of examining effects of meridional amblyopia. No important changes to eligibility criteria were adopted after trial commencement.

Eligible patients were randomized into two groups, which corresponded roughly to each other in terms of age composition and type of ophthalmopathologic disorder of their visual acuity, i.e. the expression of their respective amblyopia. The patients were allocated to the both groups in full randomization at 1:1 ratio without any restrictions, e.g., such as blocking.

The first group consisted of 17 children (34 eyes) aged 10 to 13 (average 11.6 ± 0.3) years. Of these, 6 patients had myopic astigmatism, 9 patients hypermetropic astigmatism and 2 patients mixed astigmatism. Refractive amblyopia (only against the background of astigmatism) was found in 4 patients, in combination with dysbinocular amblyopia (against the background of strabismus) in 4 patients and in combination with a pathology of the eye background in 7 patients (4 of them were additionally diagnosed with secondary strabismus). Two patients with artiphakia had a combination of refractive and obscuration-related amblyopia.

The second group consisted of 20 children (40 eyes) aged 9 to 14 (average 12.5 ±0.4) years. Of these, 12 patients had myopic astigmatism, 7 patients hypermetropic astigmatism and 1 patient mixed astigmatism. Refractive amblyopia (only against the background of astigmatism) was found in 3 patients, in combination with dysbinocular amblyopia (against the background of strabismus) in 12 patients and in combination with a pathology of the eye background in 6 patients (in 5 of them a secondary strabismus was additionally diagnosed).

A table is summarizing the main baseline demographic and clinical characteristics for the both groups taken together (Table. 1).

**Table 1:**
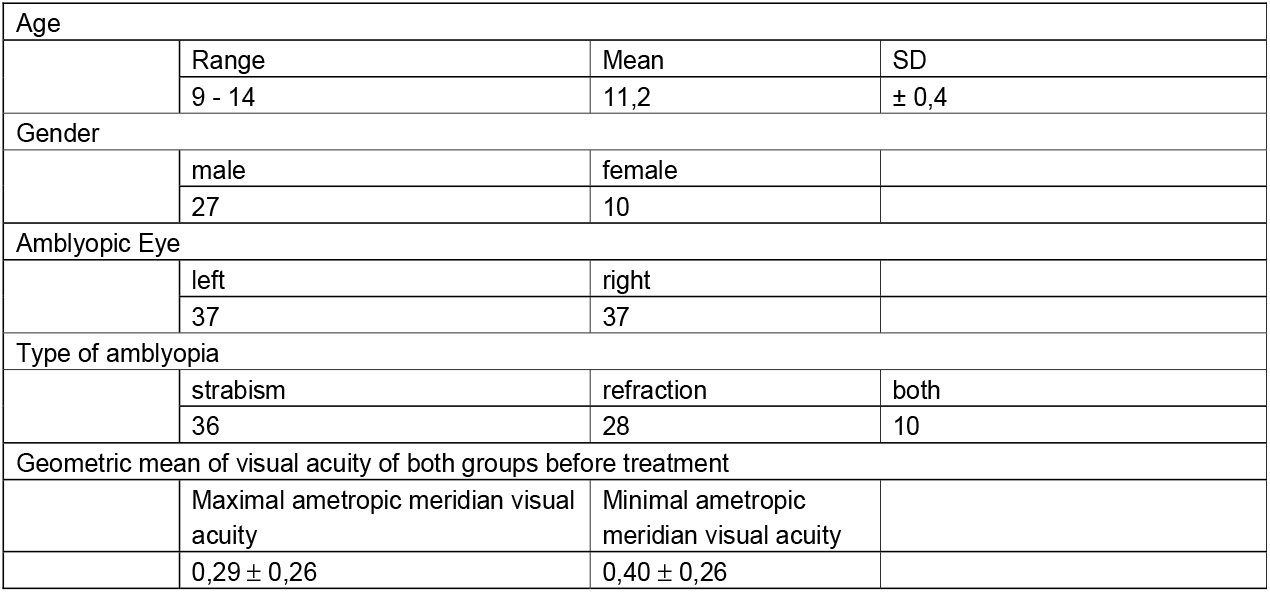
Patients’ characteristics

|  |  |  |  |
| --- | --- | --- | --- |
| Age |  |  |  |
|  | Range | Mean | SD |
| | 9 - 14 | 11,2 | $\pm 0,4$ |
| Gender |  |  |  |
|  | male | female |  |
|  | 27 | 10 |  |
| Amblyopic Eye |  |  |  |
|  | left | right |  |
|  | 37 | 37 |  |
| Type of amblyopia |  |  |  |
|  | strabism | refraction | both |
|  | 36 | 28 | 10 |
| Geometric mean of visual acuity of both groups before treatment |  |  |  |
|  | Maximal ametropic meridian visual acuity | Minimal ametropic meridian visual acuity |  |
| | $0,29 \pm 0,26$ | $0,40 \pm 0,26$ | |

### Methods

The presented study was planned and carried out as a placebo controlled, double-blinded, randomized trial in cross-over design. According to the prospective design of our study, the variation of the stimulus parameters explored was limited to the above mentioned parametric conditions that proved effective by the previous research [21, 22, 23].

For both groups’ exercises, the online vision training (Caterna Vision ltd., Potsdam, Germany; Amblyocation ltd., Liebstadt, Germany) was applied in cross-over according to the following two modifications. The first modification of the training program included a concentrically outward-moving grating, before which the computer games took place during stimulation (FAVAS = Verum). The stimulation was carried out at a spatial frequency of 0.3 cyc/deg with the temporal frequency of 1 cyc/sec, resulting in an angular velocity of 3.33 deg/sec. The second modification of the program contained the same grating, with the only difference of being exposed in a stationary (i.e., non-moving) state of presentation (Sham-FAVAS = Control). In both cases, computer games were played by the children in the foreground of screenplay, which served to bind attention in front of the stimulating grating in the background. The patients trained twice a day for 20 minutes each session. In parallel, the patients received a standard occlusion regimen.

With respect to the above described both conditions, our investigation was planned according to a two-group cross-over design (Figure. 2): The first group of patients initially completed 10 sessions using the moving sinusoidal grating in the background. After a break of one week, the same patients continued their exercises with 10 sessions this time using the stationary sinusoidal grating. In contrast, the second group of patients initially completed 10 sessions with the stationary gratings. After a break of one week, the same patients continued their exercises with 10 sessions this time using the moving sinusoidal grating. All the patients performed in the respective 10 sessions of visual exercises monocularly for 10 minutes for each eye separately.

**Figure 2:**
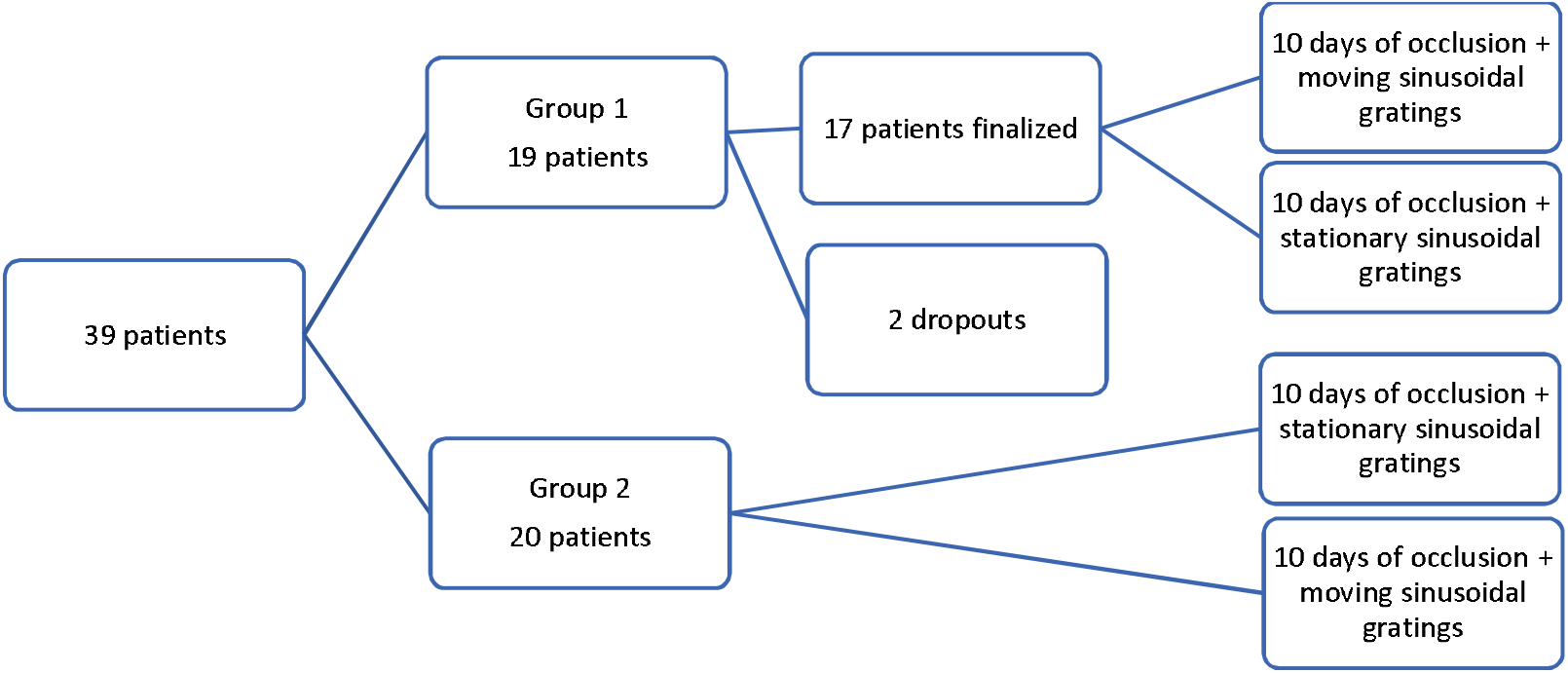
Patients’ allocation over trial conditions

In accord with our objectives, we examined our patients with regard to their best-corrected visual acuity using a meridionally direction-sensitive visual test inventory especially developed at the Kharkevich Institute [42]. Thereby we estimated the monocular corrected visual acuity in all patients along 4 meridians: 0°, 45°, 90° and 135°. The tests with these visual examinations were carried out before and after the respective treatment series of our functional amblyopia therapy.

## Results

As a result, we plot the mean visual acuity for the both groups of patients at the baseline, after the initially performed training series and after cross over (Figure. 3). Different graphs show effects for the most ametropic meridian vs least ametropic meridian. Additionally, Table. 2 is showing the graphically presented results in their digital representation.

**Figure 3:**
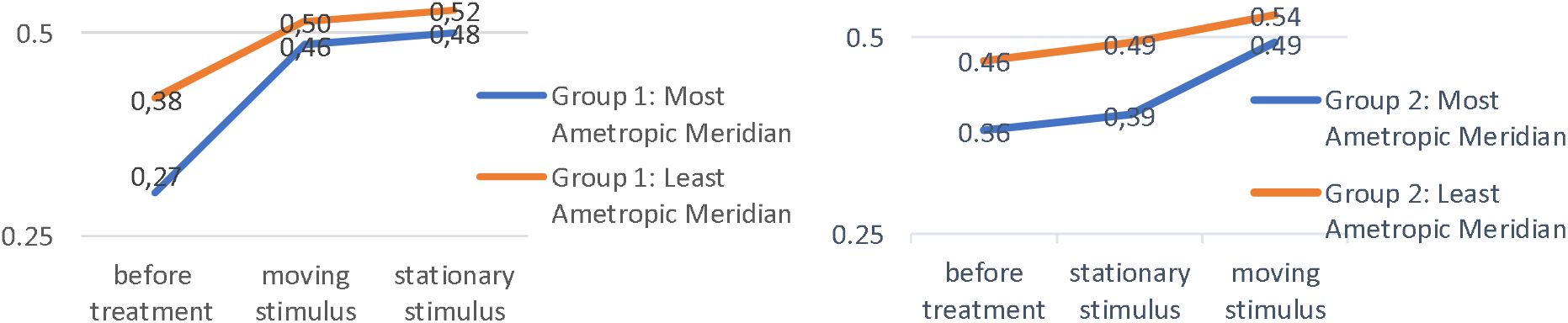
Mean decimal visual acuity results for baseline and treatment before vs. after cross over

**Table 2:**
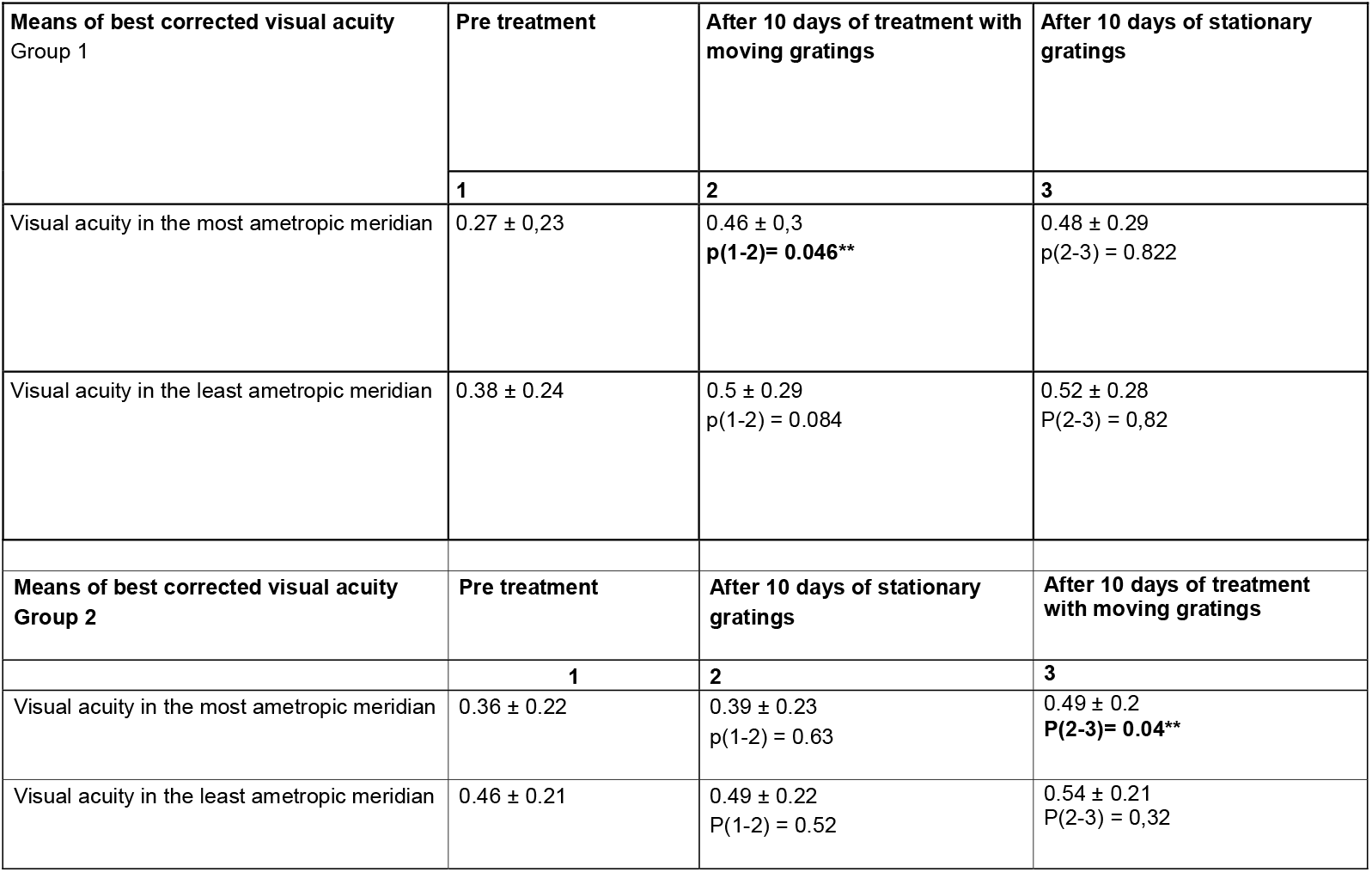
Mean visual acuity results for the both groups of patients. Significance revealed by independent t-Test is marked by double asterisk (p<0.05).

| Means of best corrected visual acuity Group 1 | Pre treatment | After 10 days of treatment with moving gratings | After 10 days of stationary gratings |
| --- | --- | --- | --- |
|  | 1 | 2 | 3 |
| Visual acuity in the most ametropic meridian | 0.27 ± 0.23 | 0.46 ± 0.3<br>p(1-2)= 0.046** | 0.48 ± 0.29<br>p(2-3) = 0.822 |
| Visual acuity in the least ametropic meridian | 0.38 ± 0.24 | 0.5 ± 0.29<br>p(1-2) = 0.084 | 0.52 ± 0.28<br>P(2-3) = 0.82 |
| Means of best corrected visual acuity Group 2 | Pre treatment | After 10 days of stationary gratings | After 10 days of treatment with moving gratings |
|  | 1 | 2 | 3 |
| Visual acuity in the most ametropic meridian | 0.36 ± 0.22 | 0.39 ± 0.23<br>p(1-2) = 0.63 | 0.49 ± 0.2<br>P(2-3)= 0.04** |
| Visual acuity in the least ametropic meridian | 0.46 ± 0.21 | 0.49 ± 0.22<br>P(1-2) = 0.52 | 0.54 ± 0.21<br>P(2-3) = 0.32 |

In the measurements of the corrected visual acuity along four meridians, a statistically significant improvement was found with alignment of the directional optical characters close to the meridian with maximal ametropia, and the minimal improvement in the orientation perpendicular thereto. In the patients of the both groups, the corrected visual acuity had significantly increased as a result of the treatment performed in the stage with a series of exercises with the moving sinusoidal grating. After the stage of treatment using the stationary grating, however, there was found no statistically significant improvement.

For our significant binary outcomes, both absolute and relative effect sizes were calculated Absolute Cohen’s d = 2,40 with a correlation coefficient r of 0.77 for the most ametropic meridian for Group 1 with moving gratings. Absolute Cohen’s d = 1,40 with a correlation coefficient r of 0,57 for the most ametropic meridian for Group 2 with moving gratings.

## Discussion

This is the first study with cross-over design to investigate whether FAVAS is more effective as a complementary treatment for patching than patching alone. Visual training of patients with meridional amblyopia by a series of online exercises using special computer games which contained moving gratings (FAVAS) resulted in a statistically significant positive dynamics of visual acuity development in the most ametropic meridian as compared to the least ametropic meridian, including cases with complex disturbed refraction and/or pathologies of their eye background.

Thus, the significant improvements were found in the meridian with maximum ametropia. Significantly less effects were found in the meridian with minimum ametropia. At the same time, no such improvement was achieved in the cross-over design after the respective analogue exercise series of sham FAVAS with stationary gratings.

Home-based therapeutic FAVAS computer games are shown to be effective in the presented as well as in the reported earlier trials [21, 22, 23, 30, 35] and therefore promising to overcome the shortcomings of stand-alone occlusion therapy. Accordingly, they are meanwhile reimbursed by selected insurance companies in Germany as a second-line therapy, in the way they are offered today.

It has been under lively debate whether in the therapeutic value of these offers, the computer games per se or the stimulating background with moving sinusoidal gratings were the main reason for the visual improvement per se [5] and whether stimulating approaches should be used for cases of therapy resistant amblyopia only or should they be firstly prescribed from the early beginning as a treatment to newly identified patients as well. With regard to the criticism of Bau et al [5], however, it must be said that their occlusion scheme adopted from Campbell of occluding only within the practice period (minimal occlusion) does not represent the objective of the combination therapy of FAVAS together with occlusion *lege artis* and has never been proposed to be applied this minimal way.

Additionally, the results of our trial strongly demonstrate that the sensory-motor interaction in the computer-gaming activities alone cannot be the main reason for the treatment-induced vision enhancement since the visual-acuity gain was only obtained in the experimental condition with the moving FAVAS background stimulus grating, but not in the control condition with the stationary background. This is further underlined by the fact that the maximal gains were selectively addressing the highest refractive meridians and less affecting the lowest refractive ones. Furthermore, these effects are in no means limited to the specificities of meridional amblyopia alone, since a comparable gain was shown in the earlier trials with other forms of refractive and strabismic amblyopia as well [21, 22, 23].

Discussing these significant effects of moving versus stationary stimulus gratings in comparison to the results of Campbell et al. [9], it must be premised, that the prototype for our design of the computer-based FAVAS background stimulation was not in as much the above discussed Cambridge Stimulator per se, but rather a mechanical repetitive-grating arrangement proposed by Haase [17] and Osterloh [36]. This arrangement was inspired by a visuo-motor and/or visuo-sensor pleoptic stimulation approach which had been originally developed by Otto and Stangler [38]. Initially, the latter authors described and confirmed in their further related research a beneficial effect of moving light stripe application on the fixation stability of amblyopic eyes [36, 37, 46]. Additionally, Haase [17] and Osterloh [36] found an amplification of this effect using, instead of single bars, a whole repetitively displayed grating of slightly blurred moving light stripes as a background stimulus in a fixation task.

As a hypothesis, the enhanced treatment effect caused by the spatio-temporal periodicity of the stimulus grating itself is supposed to be achieved in that the induced optomotoric and optosensoric stimulus effects mutually interact. This is possibly due to a co-operative synergy in the filter characteristics of two systems of different sensory transmission channels [8, 29]. These channels are shown to correspond, roughly speaking, to the neuronal correlates of focal processing of visually perceived form and configurative detail (sustained channels, parvocellular system) versus the ambient processing of motion and rapid change (transient channels, magnocellular system). The filters of each of these channels are selectively tuned to a narrow band of spatial frequencies [7, 10], associated each, as has been found by later research, additionally with an appropriate temporal frequency range constrained to one another in reciprocal, i.e., inversely counterbalanced synergy [24, 25].

This interdependence may be due to a kind of non-linear coupling in the synchronisation of brain functioning by means of so-called neuronal synfire-chains [1, 2]. According to this view, the drifting light stripe stimulation under spatial and temporal periodicity of a limited frequency bandwidth (which characterizes the spiral pattern of the pleoptic centrophor exercises by Bangerter [4] und Cüppers [11] too) is applied to the disturbed system’s dynamics as an external order parameter. Thus, it is expected to affect the internal coherence of normally highly ordered cortical processing states which were shown to be desynchronised, however, in amblyopia [14, 26, 41]. Additionally, to the originally reported fixation-stabilizing optomotoric effect of the light stripe stimulation, we suppose the drifting sinusoidal grating to induce an optosensoric resonance in complementary bandwidth selective filters of form vs. motion channel systems due to their cooperative visual processing. According to our preliminary hypothesis, the internal synchronisation in the focal form channels is externally supported via resonant phase coupling to ambient motion channels [22, 23].

A treatment sharing some common theoretical fundamentals with the spatial-frequency approach of Campbell’s as opposed to our proposed stimulation has been developed and verified in placebo-controlled trials by Polat et. al. [39]. In their “perceptual-learning” training, high-contrast grating stimuli in a collineated arrangement of Gabor patches are used as peripheral flankers for the detection enhancement of low-contrast central Gabor patches of common spatial frequency. Their studies demonstrated a considerable efficacy of such training for the improvement of contrast sensitivity and visual acuity in adult amblyopic patients. In common with Campbell’s and our approaches, Polat’s training is also based on the assumption of an ambient cooperative effect of peripheral stimulation on macular low-level focal visual processing. However, similar to Campbell’s and differing from our proposed stimulus arrangement, in Polat’s setting the peripheral influence on central vision was mediated by stationary, not by moving stimuli, thus, an influence of time frequency is excluded. Additionally, differing from Campbells and our approaches, the training is based on a detection task with respect to the foveal Gabor stimulus, which might be easily coped with by adults, but might be tedious to children.

## Conclusion

Confirmatory, our results show the primary endpoint of two lines of visual acuity increase in less than 10 days of primary combination of FAVAS and occlusion. The higher effect size in the verum condition of group 1 (Cohen’s d = 2.4) compared to group 2 (Cohen’s d = 1.2) means that an initial combination therapy of occlusion and FAVAS in the sense of a first-line therapy is superior to a second-line therapy.

Our conclusion of the present results is that Focal Ambient Visual Acuity Stimulation (FAVAS) in combination with occlusion is more effective than occlusion alone and should be applied not only in therapy-resistant cases, but also in all cases of initial amblyopia treatment as early as possible.

## Data Availability

All data is on file and available frome the corresponding author.

## Notes

### Competing Interest Statement

Uwe Kaempf is scientific consultant of Caterna Vision GmbH and Shareholder of Amblyocation GmbH

### Clinical Trial

DRKS00022791

### Funding Statement

The funding for this study is provided from the Russian Academy of Sciences (Kharkevich Institute).

### Author Declarations

Russian Academy of Sciences Kharkevich Institute Scientific Council Bolshoy Karetny per. 19 127051 Moscow Russian Federation

